# Self-reported diagnoses as training labels for finding uncoded chronic fatigue, fibromyalgia, and endometriosis in the All of Us Research Program

**DOI:** 10.64898/2026.10.19.26365869

**Authors:** Amir Asiaee, Kaveh Aryan

## Abstract

**Objective:** Case-finding models for underdiagnosed conditions are usually trained on diagnosis codes, which are missing from the records of many patients with a clinical diagnosis. We asked whether models trained on self-reported diagnoses rank uncoded self-reporters higher than models trained on later codes, and how a score fit only from released summary counts compares with a participant-level model.

**Methods:** In the All of Us Research Program, we followed survey respondents with no qualifying diagnosis code by the survey date for later codes. For chronic fatigue, fibromyalgia, and endometriosis (participants recorded as female at birth), we fit a naive Bayes score with empirical-Bayes shrinkage, from released feature-by-label counts for the self-reported diagnosis and from exact counts inside the data enclave for a later code, and an elastic net on participant-level data for each label. In a random 20% validation split, we measured discrimination of self-report, of uncoded self-reporters (no qualifying code observed), and of later codes.

**Results:** For chronic fatigue (149,860 respondents, including 7,770 self-reporters, 95% with no code observed), the summary-count score trained on self-report ranked uncoded self-reporters with an area under the receiver operating characteristic curve (AUROC) of 0.74 (95% CI, 0.72– 0.75), against 0.67 (95% CI, 0.65–0.69) when trained on later codes. For later codes, the two scores were similar (0.69 vs 0.70). Fibromyalgia showed the same pattern (0.73 vs 0.64 and 0.74 vs 0.72). Elastic nets trained on self-report ranked uncoded self-reporters higher than the summary-count scores (0.80 vs 0.74 and 0.78 vs 0.73) but discriminated later codes less well than code-trained elastic nets. For endometriosis, the self-report score also ranked uncoded self-reporters above the code-trained score (0.61 vs 0.53), but discrimination was low and it did not discriminate later codes (0.50 vs 0.70).

**Conclusion:** The choice of training label changes which patients a case-finding model ranks highly. For all 3 conditions, self-reported diagnoses were the more useful label for finding uncoded self-reporters. For chronic fatigue and fibromyalgia, the self-report score also discriminated later codes as well as the code-trained score. Independent clinical validation is needed before these models guide case finding.

## 1 Introduction

A case-finding model learns whatever its training label encodes. When a condition is coded whenever it is diagnosed, a diagnosis code is a reasonable label. When a condition is often diagnosed without being coded, or diagnosed outside the health system that holds the record, a model trained on codes learns which patients reach a code: which clinicians code it, which patients see specialists, and which sites use which codes. We call this the label problem of case finding.

Positive-unlabeled (PU) learning states the problem formally. Let *Y* denote disease, *L* an observed label such as a code, and *X* chart features. If labels have no false positives, *P* (*L* = 1 | *Y* =0, *X*) = 0, and labeling is independent of the features among true cases, *P* (*L* = 1 | *Y* = 1, *X*) = *c* for a constant *c* (selected completely at random), then *P* (*L* = 1 | *X*) = *c P* (*Y* = 1 | *X*), and a model of the label ranks patients as a model of disease would [1, 2]. When labeling depends on the features, the label model inherits that dependence [3]. Coding plausibly depends on features such as visit volume, specialist care, and site.

Informatics work on noisy labels has mostly improved how codes are used rather than replacing them. PheNorm normalizes counts of a condition’s codes and mentions into a phenotype score without chart review [4]. Weakly semi-supervised methods combine such surrogates with a small set of chart-reviewed labels [5], and SCANER calibrates risk estimates when event timing is recorded with error [6]. For Long COVID, several models use the U09.9 code or clinic attendance as the label [7–9], and LATCH refines code-based learning with chart-reviewed reference labels [10]. These approaches need either the code to be a reasonable surrogate or chart review to correct it. Neither holds well for conditions that are rarely coded and rarely reviewed.

In a companion study in the same base population, we measured how often a self-reported diagnosis carries a code [11]. Among respondents with at least 5 years of records who reported current care for the condition, the share coded relative to well-coded anchor conditions was 0.28 for chronic fatigue, 0.49 for endometriosis, and 0.73 for fibromyalgia, so coding is incomplete among people who report a diagnosis, by amounts that differ by condition. Patients’ own reports of a diagnosis are a different surrogate: they do not depend on whether the diagnosing clinician coded it.

In this paper, we compare the two surrogates as training labels for 3 conditions. We asked (1) whether models trained on self-reported diagnoses rank self-reporters with no qualifying code (uncoded self-reporters) higher than models trained on later codes, (2) how the two labels compare in discriminating later codes, the usual training label, and (3) how a score fit only from released summary counts, which can leave a data enclave, compares with a participant-level model. Our contribution is an empirical comparison of training labels under the constraints of a research data enclave, not a new learning algorithm.

Statement of significance.

**Statement of significance.**
|  |  |
| --- | --- |
| Problem | Case-finding models are trained on diagnosis codes, which are missing from the records of many patients with a clinical diagnosis. |
| What is already known | Codes are noisy surrogates, and existing methods refine code-based labels or correct them with chart review. |
| What this paper adds | Models trained on self-reported diagnoses ranked uncoded self-reporters higher than code-trained models for all 3 conditions, though discrimination was low for endometriosis. A summary-count version can be exported. |
| Who would benefit | Teams building case-finding tools for inconsistently coded conditions, especially inside data enclaves. |

## 2 Materials and Methods

### 2.1 Data source and population

Figure 1 summarizes the design, from the base population to the exported models. We used the All of Us Research Program Registered Tier Dataset v9 (curated data repository R2025Q4R6, data cutoff January 1, 2025) [12], in which dates are shifted backward by 1 to 365 days per participant, preserving intervals within a person. The base population, shared with the companion study, was participants born in 2004 or earlier with at least 1 electronic health record (EHR) visit both from July 1, 2021, to June 30, 2022, and from July 1, 2022, to June 30, 2023. Day 0 was each participant’s earliest completion date of the Personal and Family Health History or Personal Medical History survey. For each condition, the analysis cohort was participants aged 18 years or older at day 0 (calendar year minus birth year), with at least 1 outpatient, emergency, or inpatient visit in the 365 days before day 0, and with no qualifying code for the condition on or before day 0. Sex was the participant-reported sex at birth in the program record. We restricted the endometriosis cohort to participants recorded as female, and sex subgroups used the female and male categories. Because the base population required visits in 2021–2023, which can fall after day 0 for participants surveyed earlier, the cohorts are conditioned on later contact with care.

**Figure 1:**
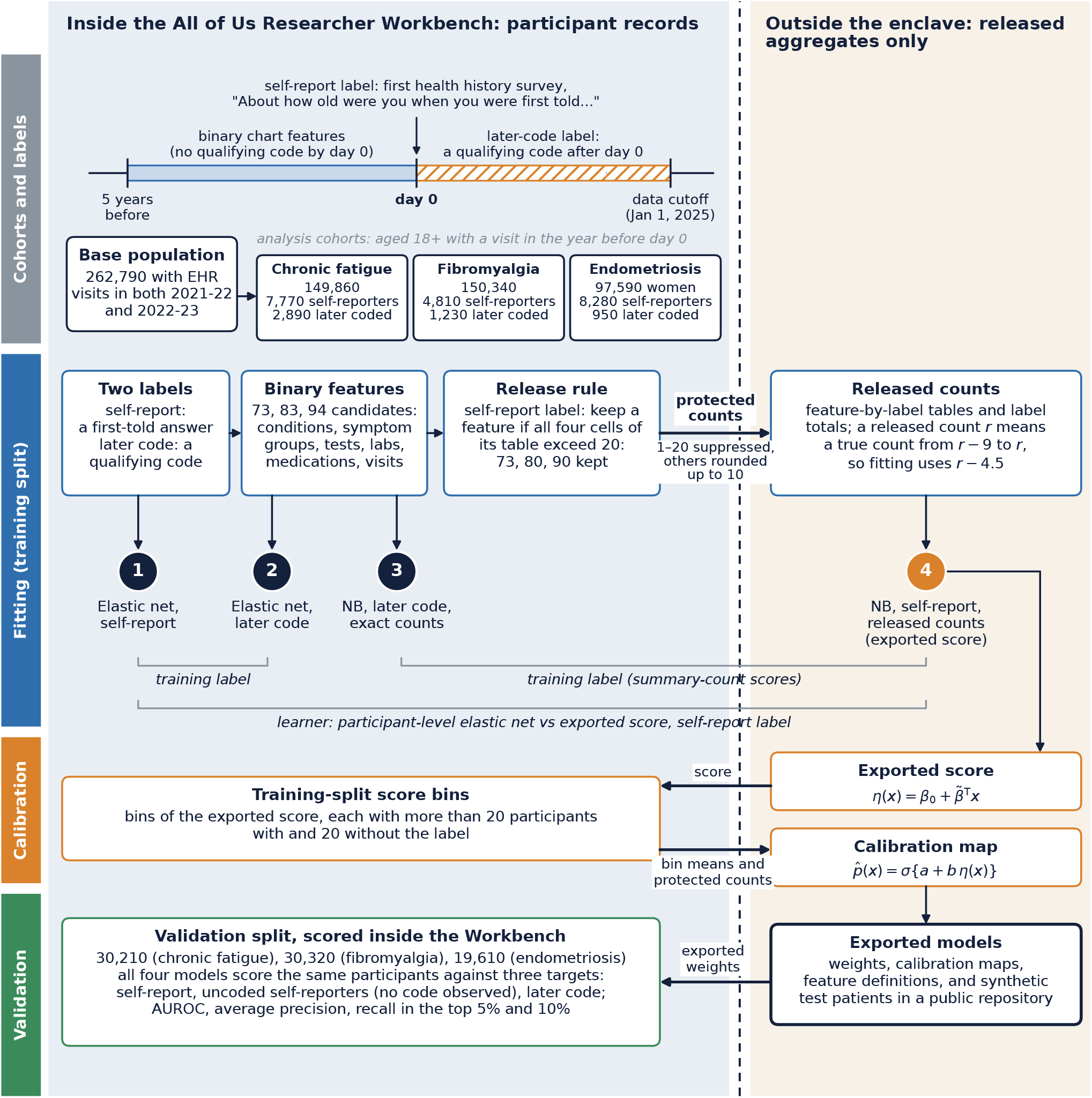
Study design inside and outside the All of Us enclave. Day 0 is each participant’s first health history survey. Chart features come from the 5 years before it, the self-report label from the survey, and the later-code label from codes recorded after it. For each condition, four models were fitted to the same training participants: models 1 to 3 inside the enclave and model 4, the exported score, outside it from released counts of the features eligible for the self-report label. Brackets mark the comparisons in Table 1. Arrows across the dashed line are the only transfers between the enclave and the outside: protected counts and calibration-bin summaries leave it, and the exported score and its weights return to it for scoring. Counts are rounded up to the next multiple of 10, and cohort totals are sums of the rounded training and validation counts.

### 2.2 Labels and evaluation targets

The self-report label was any recorded answer, including a skipped answer, to the survey item “About how old were you when you were first told you had [condition]?” The survey shows this item only to participants who report the condition, and we treated participants without such an answer as not reporting it. We took the answer from any completed survey response, not only the response dated day 0. The later-code label was any qualifying International Classification of Diseases, Tenth Revision, Clinical Modification (ICD-10-CM) code recorded after day 0 in the condition table: G93.3 or R53.82 (chronic fatigue), M79.7 (fibromyalgia), or N80 (endometriosis), which we matched by prefix on source concepts. We observed codes through the data cutoff, so the follow-up time after day 0 varied by participant, and “no code observed” means no qualifying code in this dataset, not that a diagnosis was never recorded anywhere.

**Table 1:** Discrimination by training label: self-report versus later codes. Validation split (random 20%). We rounded counts up to a multiple of 10 and limited the endometriosis cohort to participants recorded as female at birth. Summary-count scores are naive Bayes with empirical-Bayes shrinkage (we fit the self-report score from released counts, so it can be exported). We fit elastic nets to participant-level data. Uncoded: self-reporters with no qualifying code observed after the survey (negatives are all other validation participants). Later code: a qualifying code recorded after the survey. Recall: percentage of all uncoded self-reporters ranked within the highest-scoring 5% or 10% of the full validation split. AUROC, area under the receiver operating characteristic curve, with 95% bootstrap confidence intervals in parentheses.

| Model | Trained on | AUROC, self-report |  | Recall, uncoded, % |  | AUROC, later code |
| --- | --- | --- | --- | --- | --- | --- |
|  |  | All | Uncoded | Top 5% | Top 10% |  |
| *Chronic fatigue*: 30,210 validation participants, 1,620 self-reporters, 1,540 uncoded |  |  |  |  |  |  |
| Summary counts | Self-report | 0.74 (0.73–0.76) | 0.74 (0.72–0.75) | 22 | 33 | 0.69 (0.67–0.71) |
| Summary counts | Later code | 0.68 (0.66–0.69) | 0.67 (0.65–0.69) | 19 | 29 | 0.70 (0.68–0.72) |
| Elastic net | Self-report | 0.80 (0.79–0.81) | 0.80 (0.79–0.81) | 31 | 45 | 0.60 (0.58–0.63) |
| Elastic net | Later code | 0.66 (0.65–0.68) | 0.65 (0.64–0.67) | 18 | 28 | 0.72 (0.70–0.74) |
| *Fibromyalgia*: 30,320 validation participants, 930 self-reporters, 840 uncoded |  |  |  |  |  |  |
| Summary counts | Self-report | 0.72 (0.71–0.74) | 0.73 (0.71–0.74) | 20 | 33 | 0.74 (0.70–0.77) |
| Summary counts | Later code | 0.64 (0.63–0.66) | 0.64 (0.62–0.66) | 11 | 19 | 0.72 (0.69–0.76) |
| Elastic net | Self-report | 0.78 (0.77–0.80) | 0.78 (0.77–0.80) | 31 | 44 | 0.73 (0.69–0.76) |
| Elastic net | Later code | 0.72 (0.71–0.74) | 0.72 (0.70–0.73) | 17 | 27 | 0.80 (0.77–0.82) |
| *Endometriosis* (female participants): 19,610 validation participants, 1,640 self-reporters, 1,580 uncoded |  |  |  |  |  |  |
| Summary counts | Self-report | 0.61 (0.59–0.62) | 0.61 (0.60–0.62) | 8 | 15 | 0.50 (0.45–0.54) |
| Summary counts | Later code | 0.53 (0.52–0.54) | 0.53 (0.51–0.54) | 5 | 11 | 0.70 (0.66–0.73) |
| Elastic net | Self-report | 0.64 (0.62–0.65) | 0.64 (0.63–0.65) | 11 | 20 | 0.38 (0.34–0.43) |
| Elastic net | Later code | 0.47 (0.46–0.49) | 0.46 (0.45–0.48) | 5 | 9 | 0.77 (0.74–0.80) |

We evaluated models against 3 targets in the validation split: (1) self-report, (2) uncoded self-reporters, and (3) a later code. For target 2, positives were self-reporters with no qualifying code observed after day 0, and negatives were all other validation participants, including self-reporters who were later coded. Because we still defined target 2 by self-report, it shows which participants a model ranks highly among those a code-trained model cannot learn from. It is not an independent clinical reference standard.

### 2.3 Features

Predictors were binary indicators of chart events in the 5 years before day 0 (or the 365 days before, for recent features): age band and sex, symptom and related-condition codes, counts of symptom groups, procedures and evaluations, laboratory tests with a recorded measurement, medication exposure records, and health care use. Each condition had its own registry of codes (ICD-10-CM, Current Procedural Terminology and Healthcare Common Procedure Coding System, LOINC, and RxNorm), including condition-specific features such as pelvic pain, dysmenorrhea, pelvic ultrasound, and hormonal therapy for endometriosis (Supplementary Tables S1–S3). The label’s own codes were never predictors. ICD-10-CM predictors came from both the condition and observation tables, because the All of Us data model files some diagnosis codes as observations. The labels used the condition table only. A chart feature was 0 when no qualifying record existed, which does not establish that the finding was clinically absent.

### 2.4 Models

Models trained on participant-level data cannot leave the All of Us Researcher Workbench. We therefore built the exportable score from feature-by-label tables in a random 80% training split. A feature was eligible if all 4 cells of its 2 *×* 2 table with the label exceeded 20. For the self-report label, 73 of 73 candidate features were eligible for chronic fatigue, 80 of 83 for fibromyalgia, and 90 of 94 for endometriosis. Because we rounded each released cell up to a multiple of 10, we subtracted the expected rounding excess (4.5) from every cell before fitting. For feature *j*, let 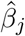be the log odds ratio between the feature and the label and *s*_*j*_ its standard error. We shrank each 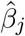toward 0 under a normal prior,

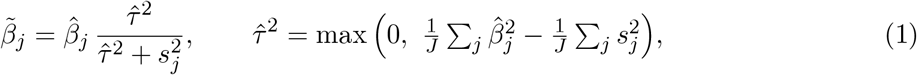

and a participant’s score was 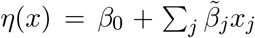, with 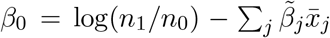 where *n*_1_ and *n*_0_ are the numbers of label-positive and label-negative training participants and 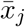 is the feature’s prevalence. This is a naive Bayes score built from shrunken marginal log odds ratios. Supplementary Methods derives it from the naive Bayes model and gives every computation, from the released tables and their rounding to the variance and shrinkage estimates, the intercept,the calibration bins and map, and the elastic-net objective. The released tables are sufficient to reconstruct it for the eligible features. Suppression and rounding can change it relative to a fit from exact counts. We mapped scores to probabilities of the label by logistic regression on binned training-split scores, each bin with more than 20 label-positive and 20 label-negative participants.

The comparison score trained on later codes used the same construction from exact counts inside the Workbench, and we determined eligibility separately for that label. We fit elastic nets to participant-level training data for each label (standardized features, logistic loss, mixing parameter 0.5, inverse penalty strength 0.05, and the SAGA solver in scikit-learn), with no tuning and no class weighting. We ran the analyses in the Researcher Workbench with Python 3.12 with NumPy 2.4, pandas 2.3, and scikit-learn 1.8.

### 2.5 Evaluation

In the validation split, we report the area under the receiver operating characteristic curve (AU-ROC) with 95% confidence intervals (CIs) from 200 participant-level bootstrap resamples of the validation split, with the fitted models held fixed (percentile intervals). We also report the average precision, compared with each target’s prevalence, and recall among the 5% and 10% highest-scoring validation participants. Recall is the percentage of all uncoded self-reporters ranked within that share of the full validation split, counting exactly 5% or 10% of participants. We estimated differences between the self-report-trained and code-trained summary scores with paired percentile intervals from 200 bootstrap resamples, each scoring both models on the same resampled participants. These paired comparisons counted participants at or above the corresponding score quantile, which can include a few more participants when scores are tied. Intervals reflect sampling of validation participants only, not model refitting or variation across health systems. Sensitivity analyses repeated the comparison on a second 80/20 split, defined by a hash of the participant identifier, with every model refit. They changed the target, the timing of both labels, and the feature set one at a time, added two simple baselines, and examined subgroups by sex, age, and survey period. We also assessed the calibration of the exported scores in the validation split of Table 1 (Supplementary Methods, which also defines each measure). We assessed reporting against TRIPOD+AI [13] (checklist in Supplementary Table S4).

### 2.6 Disclosure control and ethics

We ran all analyses of participant data inside the Researcher Workbench and exported only aggregate tables and performance metrics. We suppressed counts of 1 to 20, rounded other counts up to a multiple of 10, and, within each table, suppressed a further count wherever a suppressed count could otherwise be recovered from the table’s totals (complementary suppression). We withheld recall and precision at a ranking threshold when the number of target participants among those flagged was 1 to 20. We checked the numbers reported in this article and its supplement so that no count of 1 to 20 can be derived from them. Percentages from the training-split feature tables, which we reconstructed from the rounded counts, are approximate.

All of Us participants provide informed consent at enrollment under a protocol overseen by the program’s institutional review board. We analyzed de-identified Registered Tier data as authorized researchers [12].

## 3 Results

### 3.1 Cohorts

The chronic fatigue cohort had 149,860 respondents, of whom 7,770 (5.2%) reported chronic fatigue. Of these self-reporters, 7,350 (95%) had no qualifying code observed after the survey, and 2,450 of the 2,890 participants with a later code (85%) had not reported the condition. The fibromyalgia cohort had 150,340 respondents: 4,810 (3.2%) reported fibromyalgia, 4,270 of them (89%) had no code observed, and 680 of the 1,230 with a later code (55%) had not reported it. The endometriosis cohort had 97,590 participants recorded as female: 8,280 (8.5%) reported endometriosis, 8,020 of them (97%) had no code observed, and 680 of the 950 with a later code (72%) had not reported it. Totals are sums of separately rounded training and validation counts. Supplementary Table S5 gives the participant flow.

### 3.2 Discrimination by training label

For chronic fatigue, the summary-count score trained on self-report discriminated self-reporters with an AUROC of 0.74 (95% CI, 0.73–0.76) and uncoded self-reporters with 0.74 (95% CI, 0.72–0.75), and ranked 22% of uncoded self-reporters within its top 5% of scores and 33% within its top 10% (Table 1). Trained on later codes, the same construction reached 0.67 (95% CI, 0.65–0.69) for uncoded self-reporters and ranked 19% and 29% of them within the top 5% and 10%. The paired differences in recall were 3.3 (95% CI, 2.2–4.4) and 4.6 (95% CI, 3.4–5.8) percentage points, and the AUROC difference for all self-report was 0.063 (95% CI, 0.059–0.068). For later codes, the two summary scores had similar AUROCs (0.69 and 0.70). The participant-level elastic nets showed the same label contrast for uncoded self-reporters (0.80 vs 0.65), and the self-report-trained elastic net ranked them higher than the summary-count score trained on the same label (0.80 vs 0.74). For later codes, however, it was lower than the code-trained elastic net (0.60 vs 0.72).

For fibromyalgia, the self-report score ranked uncoded self-reporters with an AUROC of 0.73 (95% CI, 0.71–0.74), with 20% and 33% of them within its top 5% and 10%. The code-trained score reached 0.64 (95% CI, 0.62–0.66), 11%, and 19%. The paired differences in recall were 8.5 (95% CI, 6.2–10.6) and 14.1 (95% CI, 11.4–16.7) percentage points, and the AUROC difference for all self-report was 0.083 (95% CI, 0.074–0.093). The self-report-trained elastic net reached 0.78 for uncoded self-reporters. For later codes, the summary scores were similar (0.74 [95% CI, 0.70–0.77] and 0.72 [95% CI, 0.69–0.76]), while the code-trained elastic net was higher than the self-report-trained one (0.80 vs 0.73).

For endometriosis, the self-report score again ranked uncoded self-reporters higher than the code-trained score, although both AUROCs were low (0.61 [95% CI, 0.60–0.62] vs 0.53 [95% CI, 0.51–0.54]). Recall within the top 5% was 8.3% vs 5.3%, a difference of 3.0 (95% CI, 2.0–4.4) percentage points, and the self-report-trained elastic net reached 0.64. The self-report score did not discriminate later codes (0.50 [95% CI, 0.45–0.54], against 0.70 [95% CI, 0.66–0.73] for the code-trained score, and 0.38 vs 0.77 for the elastic nets).

Average precision for uncoded self-reporters was 0.16 for chronic fatigue, 0.08 for fibromyalgia, and 0.11 for endometriosis with the self-report score, against target prevalences in the validation split of about 5.1%, 2.8%, and 8.1%. For self-report as a whole, the AUROC of the self-report score was 0.73 among participants recorded as female and 0.75 among those recorded as male for chronic fatigue, and 0.69 and 0.70 for fibromyalgia. Supplementary Table S7 gives subgroup AUROCs with intervals for uncoded self-reporters.

### 3.3 Features associated with self-report

In the training split (feature definitions in Supplementary Tables S1–S3), the largest chronic fatigue weights were for mast cell disorder, fibromyalgia (recorded for about 19% of self-reporters and 2% of other respondents), Ehlers-Danlos syndrome or hypermobility, a medication used for postural tachycardia, a tilt-table test, and a stimulant or wakefulness drug. Fatigue codes were recorded for about 27% and 16%. The largest fibromyalgia weights were for milnacipran exposure (about 1.5% vs under 0.1%), Ehlers-Danlos syndrome or hypermobility, female sex (87% vs 62%), a medication used for postural tachycardia, muscle relaxants (29% vs 11%), gabapentinoids (44% vs 19%), stimulants, an anti-cyclic citrullinated peptide (anti-CCP) antibody test, selective serotonin or serotonin-norepinephrine reuptake inhibitors, and a rheumatoid factor test.

Endometriosis self-reporters were older than other participants (about 51% vs 44% aged 60 years or older, and 11% vs 22% aged 18 to 39 years) and less often had hormonal contraceptive or progestin exposure (11% vs 17%) or codes for heavy or irregular menstrual bleeding (3.6% vs 6.5%) in the 5 years before the survey. Pelvic pain (9% vs 7%), dysmenorrhea, and pelvic imaging differed little. The largest positive weights were for fibromyalgia, interstitial cystitis, mast cell disorder, gonadotropin-releasing hormone agonist or antagonist therapy, Ehlers-Danlos syndrome or hypermobility, a medication used for postural tachycardia, gabapentinoids, and a tilt-table test. We did not assess disease activity or reasons for later coding.

### 3.4 Sensitivity analyses

On the second split, AUROCs for self-report and for uncoded self-reporters differed by 0.03 or less from Table 1, and those for later codes, which rest on fewer events, by up to 0.06 (Supplementary Table S6). In every check, the self-report-trained summary score ranked uncoded self-reporters above the code-trained score, by 0.01 to 0.11 in AUROC: among participants with no later code, with the self-report label limited to answers given by day 30, among participants with at least 730 days of follow-up, with both scores fitted from rounded counts of a common feature set, and in every subgroup by sex, age, and survey period (Supplementary Tables S6 and S7). It also ranked them above a logistic baseline of age, sex, and visit counts (for chronic fatigue, 0.74 vs 0.67) and above the number of symptom groups coded (0.59). Refitting the code-trained score from rounded instead of exact counts changed its AUROCs by less than 0.001. All but 20 or fewer self-reporters per condition gave their answer on day 0, and median follow-up from day 0 to the last recorded visit was 860 to 900 days. The probability maps that ship with the exported scores (Section 2.4) were close to calibrated for self-report in the validation split of Table 1, with calibration intercepts of *−*0.13 to *−*0.02 and slopes of 0.84 to 0.98 (Supplementary Methods).

## 4 Discussion

In this cohort, models trained on self-reported diagnoses ranked uncoded self-reporters higher than models trained on later codes for all 3 conditions, and the summary-count version did so with no participant-level data leaving the enclave. For endometriosis, however, discrimination was low (AUROC 0.61). For later codes, the summary scores trained on the two labels were similar for chronic fatigue and fibromyalgia (0.69 vs 0.70 and 0.74 vs 0.72) but not for endometriosis (0.50 vs 0.70), and the self-report-trained elastic nets were worse for all 3 conditions, so the label that finds uncoded self-reporters is not the label that best predicts future coding. For chronic fatigue, a later code reached about 5% of self-reporters, and 85% of participants with a later code had not reported the condition: the two labels describe largely different groups of people. The summary-count score ranked uncoded self-reporters lower than the participant-level elastic net trained on the same label for all 3 conditions (Table 1). This gap combines the learner, count rounding, and feature eligibility. Rounding mattered little: refitting the code-trained score from rounded counts changed its AUROCs by less than 0.001, and a companion Long COVID study that separated these steps on the same patients found the same for its released model [14]. For chronic fatigue every candidate feature was eligible, so the gap there reflects the learner: naive Bayes adds each feature’s marginal association with the label, whereas the elastic net fits all features jointly.

The code-trained summary score discriminated later codes better than it ranked uncoded self-reporters (AUROC 0.70 vs 0.67 for chronic fatigue and 0.72 vs 0.64 for fibromyalgia), as expected if coding depends on features of care as well as on disease. Under the PU assumptions of the Introduction, with coding independent of the features among true cases, a code-trained score would rank patients as a model of disease would and would rank uncoded cases about as well as coded ones. The two targets have different negatives, however, and uncoded self-reporters may also differ from coded patients in disease status. A self-report label avoids that dependence but brings its own: it measures a diagnosis a patient was given and remembers, not current disease [15]. Self-report is therefore a different surrogate, not a gold standard, and the uncoded-self-reporter target inherits it. Methods that combine surrogates with chart-reviewed labels [5, 10] could use self-report as one more surrogate where chart review is feasible.

An ever-told diagnosis served endometriosis poorly: discrimination was low, and the self-report score did not discriminate later codes. The survey asks whether a person was ever told they had a condition. For conditions that usually persist, such as fibromyalgia, an ever-told diagnosis plausibly indicates a condition that is still present. For endometriosis, the feature associations suggest an age-related difference between the two labels: self-reporters were older and had fewer recent menstrual and hormonal records. Among women aged 18 to 44 years, the self-report score ranked uncoded self-reporters with an AUROC of 0.63 (95% CI, 0.60–0.67), against 0.53 for the code-trained score, and it discriminated later codes (0.63, 95% CI, 0.58–0.68), which it did not in the full cohort (Supplementary Table S6). This pattern is consistent with ever-told reports by older women describing past or inactive disease, which we did not assess directly. A label should match the condition a tool is meant to find now, for example a survey item on current symptoms, recent diagnosis, or age at diagnosis.

For practice, a self-report-trained score is best used to choose whom to ask, not whom to diagnose: a flagged patient would complete a validated questionnaire before clinician review, and the score itself predicts a reported diagnosis, not disease. A companion paper describes software that delivers the exported scores to patients and clinicians through standard health information technology interfaces [16]. Responses collected through a screening app could support future model development, but symptom questionnaires and self-reported diagnoses define different outcomes and would need separate validation and an assessment of who participates.

### 4.1 Limitations

This study has several limitations. First, both labels are documentation surrogates. No independent clinical assessment established disease, so the results concern finding people who report a diagnosis absent from the coded record, not detecting undiagnosed disease. Second, we validated the models on random splits of one cohort, not at other health systems or in later periods. The 3 cohorts overlap and share preprocessing, so they are not independent replications, and the base population required visits in 2021–2023, which conditions on later contact with care. Third, the chronic fatigue survey item does not name myalgic encephalomyelitis/chronic fatigue syndrome (ME/CFS), and sex was the recorded sex at birth. We did not analyze gender. Finally, we have not validated, on real records, the mapping of these features from the research data model to the Fast Healthcare Interoperability Resources (FHIR) that a deployed tool reads.

## 5 Conclusion

In this cohort, models trained on self-reported diagnoses ranked uncoded self-reporters higher than models trained on later codes for all 3 conditions, and a version fit only from released counts retained this advantage. Only for chronic fatigue and fibromyalgia did that version also discriminate later codes as well as the code-trained score. For endometriosis, discrimination was low. The label should match the clinical question: an ever-told diagnosis served chronic fatigue and fibromyalgia better than endometriosis. Independent clinical validation is needed before such models guide case finding.

## Data Availability

The All of Us Registered Tier Dataset v9 is available to authorized researchers through the All of Us Researcher Workbench. The aggregate statistics behind every table and figure, the code and a notebook that regenerate them, and the exported chronic fatigue, fibromyalgia, and endometriosis models with their feature definitions and synthetic test patients are available at https://github.com/AsiaeeLab/unom-self-report-labels.

## Data and code availability

This study used data from the All of Us Research Program’s Registered Tier Dataset v9, available to authorized users on the Researcher Workbench. A public repository (MIT License) contains the analysis code, the aggregate statistics behind every table and figure, and the exported chronic fatigue, fibromyalgia, and endometriosis models with their feature definitions and synthetic test patients: https://github.com/AsiaeeLab/unom-self-report-labels.

## CRediT authorship contribution statement

Amir Asiaee: Conceptualization, Methodology, Formal analysis, Data curation, Writing – original draft. Kaveh Aryan: Software, Writing – review and editing.

## Acknowledgments

We gratefully acknowledge All of Us participants for their contributions, without whom this research would not have been possible. We also thank the National Institutes of Health’s All of Us Research Program for making available the participant data examined in this study. The All of Us Research Program is supported by the National Institutes of Health, Office of the Director: Regional Medical Centers: 1 OT2 OD026549; 1 OT2 OD026554; 1 OT2 OD026557; 1 OT2 OD026556; 1 OT2 OD026550; 1 OT2 OD 026552; 1 OT2 OD026553; 1 OT2 OD026548; 1 OT2 OD026551; 1 OT2 OD026555; IAA #: AOD 16037; Federally Qualified Health Centers: HHSN 263201600085U; Data and Research Center: 5 U2C OD023196; Biobank: 1 U24 OD023121; The Participant Center: U24 OD023176; Participant Technology Systems Center: 1 U24 OD023163; Communications and Engagement: 3 OT2 OD023205; 3 OT2 OD023206; and Community Partners: 1 OT2 OD025277; 3 OT2 OD025315; 1 OT2 OD025337; 1 OT2 OD025276. In addition, the All of Us Research Program would not be possible without the partnership of its participants.

## References

[1] C. Elkan and K. Noto. Learning classifiers from only positive and unlabeled data. In Proceedings of the 14th ACM SIGKDD International Conference on Knowledge Discovery and Data Mining, pages 213–220, 2008. doi: 10.1145/1401890.1401920.

[2] J. Bekker and J. Davis. Learning from positive and unlabeled data: a survey. Machine Learning, 109(4):719–760, 2020. doi: 10.1007/s10994-020-05877-5.

[3] Jessa Bekker, Pieter Robberechts, and Jesse Davis. Beyond the selected completely at random assumption for learning from positive and unlabeled data. In Machine Learning and Knowledge Discovery in Databases (ECML PKDD 2019), volume 11907 of Lecture Notes in Computer Science, pages 71–85. Springer, 2020. doi: 10.1007/978-3-030-46147-8_5.

[4] Sheng Yu, Yumeng Ma, Jessica Gronsbell, Tianrun Cai, Ashwin N. Ananthakrishnan, Vivian S. Gainer, Susanne E. Churchill, Peter Szolovits, Shawn N. Murphy, Isaac S. Kohane, Katherine P. Liao, and Tianxi Cai. Enabling phenotypic big data with PheNorm. Journal of the American Medical Informatics Association, 25(1):54–60, 2018. doi: 10.1093/jamia/ocx111.

[5] Isabelle-Emmanuella Nogues, Jun Wen, Yucong Lin, Molei Liu, Sara K. Tedeschi, Alon Geva, Tianxi Cai, and Chuan Hong. Weakly semi-supervised phenotyping using electronic health records. Journal of Biomedical Informatics, 134:104175, 2022. doi: 10.1016/j.jbi.2022.104175.

[6] Chuan Hong, Liang Liang, Qianyu Yuan, Kelly Cho, Katherine P. Liao, Michael J. Pencina, David C. Christiani, and Tianxi Cai. Semi-supervised calibration of noisy event risk (SCANER) with electronic health records. Journal of Biomedical Informatics, 144:104425, 2023. doi: 10.1016/j.jbi.2023.104425.

[7] E. R. Pfaff, A. T. Girvin, T. D. Bennett, et al. Identifying who has long COVID in the USA: a machine learning approach using N3C data. The Lancet Digital Health, 4(7):e532–e541, 2022. doi: 10.1016/S2589-7500(22)00048-6.

[8] V. M. Castro, V. Gainer, N. Wattanasin, et al. Characterization and validation of EHR computable phenotypes for Long COVID using patient-reported symptoms: insights from the nationwide RECOVER program. Journal of the American Medical Informatics Association, 33(9):1759–1769, 2026. doi: 10.1093/jamia/ocag115.

[9] C. Guardo, X. Zhang, S. Gangireddy, et al. Multi-scale data improves performance of machine learning model for long COVID identification. Communications Medicine, 6:389, 2026. doi: 10.1038/s43856-026-01621-7.

[10] C. Hong, J. Wen, H. G. Zhang, et al. Label efficient phenotyping for Long COVID using electronic health records. npj Digital Medicine, 8:405, 2025. doi: 10.1038/s41746-025-01617-y.

[11] Amir Asiaee and Kaveh Aryan. Electronic Health Record Coding of Self-Reported Diagnoses in the All of Us Research Program, 2026. Companion manuscript, submitted.

[12] The All of Us Research Program Investigators. The “All of Us” Research Program. New England Journal of Medicine, 381(7):668–676, 2019. doi: 10.1056/NEJMsr1809937.

[13] Gary S Collins, Karel G M Moons, Paula Dhiman, Richard D Riley, Andrew L Beam, Ben Van Calster, Marzyeh Ghassemi, Xiaoxuan Liu, Johannes B Reitsma, Maarten van Smeden, Anne-Laure Boulesteix, Jennifer Catherine Camaradou, Leo Anthony Celi, Spiros Denaxas, Alastair K Denniston, Ben Glocker, Robert M Golub, Hugh Harvey, Georg Heinze, Michael M Hoffman, André Pascal Kengne, Emily Lam, Naomi Lee, Elizabeth W Loder, Lena Maier-Hein, Bilal A Mateen, Melissa D McCradden, Lauren Oakden-Rayner, Johan Ordish, Richard Parnell, Sherri Rose, Karandeep Singh, Laure Wynants, and Patricia Logullo. TRIPOD+AI statement: updated guidance for reporting clinical prediction models that use regression or machine learning methods. BMJ, 385:e078378, 2024. doi: 10.1136/bmj-2023-078378.

[14] Amir Asiaee and Kaveh Aryan. Long COVID undercoding and prediction from releasable electronic health record summaries in All of Us, 2026. Companion manuscript, submitted.

[15] Brian Walitt, Robert S. Katz, Martin J. Bergman, and Frederick Wolfe. Three-quarters of persons in the US population reporting a clinical diagnosis of fibromyalgia do not satisfy fibromyalgia criteria: The 2012 National Health Interview Survey. PLOS ONE, 11(6):e0157235, June 2016. doi: 10.1371/journal.pone.0157235.

[16] Amir Asiaee and Kaveh Aryan. Design and Technical Evaluation of a Standards-Based System That Delivers Enclave-Derived Case-Finding Models to Patients and Clinicians, 2026. Companion manuscript, submitted.

