## Supplementary Material for "Self-reported diagnoses as training labels for finding uncoded chronic fatigue, fibromyalgia, and endometriosis in the All of Us Research Program"

Supplementary Methods gives the formulation of the models and of each evaluation measure, and the definitions of the sensitivity analyses. Tables S1–S3 list the candidate features of the chronic fatigue, fibromyalgia, and endometriosis models. We generated them from the same feature registries we used to fit the models. Table S4 is the TRIPOD+AI checklist. Tables S5–S7 give the participant flow and the sensitivity analyses.

### Supplementary Methods

*Released tables.* For a condition and a training label $L \in \{0, 1\}$ (self-report or a later code), let the training split contain $n_1$ participants with the label and $n_0$ without it, and let $x_{ij} \in \{0, 1\}$ be candidate feature $j = 1, \ldots, J$ of participant $i$. For each feature we counted

$$a_j = \#\{i : x_{ij} = 1, L_i = 1\}, \quad b_j = \#\{i : x_{ij} = 1, L_i = 0\}, \quad c_j = n_1 - a_j, \quad d_j = n_0 - b_j.$$

Feature $j$ is eligible when $\min(a_j, b_j, c_j, d_j) > 20$. For the self-report label, the Workbench released the four cells of every eligible feature and the totals $n_1$ and $n_0$, each rounded up to the next multiple of 10, $r = 10\lceil v/10 \rceil$, and suppressed the cells of ineligible features. A released count $r$ therefore stands for a true count $v \in \{r - 9, \ldots, r\}$. Outside the enclave we replaced each released count with the midpoint of that range, $\tilde{v} = r - 4.5$, which is the mean of $v$ when each value in the range is equally likely, and fitted the score below to $\tilde{a}_j$, $\tilde{b}_j$, $\tilde{c}_j$, $\tilde{d}_j$, $\tilde{n}_1$, and $\tilde{n}_0$. The code-trained summary score used the same construction inside the enclave from exact counts, without rounding or correction.

*Naive Bayes score.* If the features are conditionally independent given the label, with $\pi_{1j} = P(x_j = 1 \mid L = 1)$ and $\pi_{0j} = P(x_j = 1 \mid L = 0)$, the log posterior odds of the label are

$$\log \frac{P(L = 1 \mid x)}{P(L = 0 \mid x)} = \log \frac{n_1}{n_0} + \sum_j \log \frac{1 - \pi_{1j}}{1 - \pi_{0j}} + \sum_j \beta_j x_j, \qquad \beta_j = \log \frac{\pi_{1j}(1 - \pi_{0j})}{\pi_{0j}(1 - \pi_{1j})},$$

so a feature that is present adds its log odds ratio with the label. We estimated each log odds ratio from the corrected table, $\hat{\beta}_j = \log\{\tilde{a}_j \tilde{d}_j / (\tilde{b}_j \tilde{c}_j)\}$, with the large-sample variance $s_j^2 = 1/\tilde{a}_j + 1/\tilde{b}_j + 1/\tilde{c}_j + 1/\tilde{d}_j$.

*Empirical-Bayes shrinkage.* Over the $J'$ eligible features we took $\hat{\beta}_j \mid \beta_j \sim N(\beta_j, s_j^2)$ and $\beta_j \sim N(0, \tau^2)$. Then $E(\hat{\beta}_j^2) = \tau^2 + s_j^2$, which gives the moment estimate $\hat{\tau}^2 = \max\{0, J'^{-1} \sum_j (\hat{\beta}_j^2 - s_j^2)\}$. Each weight is the posterior mean $\tilde{\beta}_j = \hat{\beta}_j \, \hat{\tau}^2 / (\hat{\tau}^2 + s_j^2)$, so imprecise log odds ratios, those with a small cell, move further toward 0. The score is

$$\eta(x) = \beta_0 + \sum_j \tilde{\beta}_j x_j, \qquad \beta_0 = \log \frac{\tilde{n}_1}{\tilde{n}_0} - \sum_j \tilde{\beta}_j \bar{x}_j, \qquad \bar{x}_j = \frac{\tilde{a}_j + \tilde{b}_j}{\tilde{n}_1 + \tilde{n}_0}.$$

The intercept replaces the second sum of the posterior odds with a centering term, so that a participant with average features scores the training log odds of the label. Ranks, and therefore AUROC, average precision, and recall, do not depend on the intercept, and the calibration map below sets the probability scale.

*Calibration map.* Inside the enclave, we scored every training participant and sorted the scores. Bins were formed from the lowest score upward: a bin closed when it held more than 20 participants with the label, more than 20 without it, and at least a tenth of the training split, and any remaining participants joined the last bin. For bin $k$, the Workbench released its lowest, highest, and mean score $\bar{\eta}_k$, its number of participants $m_k$, and the number with the label $y_k$, with counts rounded up to multiples of 10. Outside, with $\tilde{m}_k = m_k - 4.5$ and $\tilde{y}_k = y_k - 4.5$, we fitted the grouped logistic model $\tilde{y}_k \sim \text{Binomial}\{\tilde{m}_k, \sigma(a + b\bar{\eta}_k)\}$, $\sigma(t) = 1/(1 + e^{-t})$, by maximizing

$$\ell(a, b) = \sum_k \Big[\tilde{y}_k(a + b\bar{\eta}_k) - \tilde{m}_k \log\{1 + \exp(a + b\bar{\eta}_k)\}\Big]$$

with Newton–Raphson iterations from $a = \log\{\sum_k \tilde{y}_k / \sum_k(\tilde{m}_k - \tilde{y}_k)\}$ and $b = 0$. The probability of the label is $\hat{p}(x) = \sigma\{a + b\,\eta(x)\}$. The model file stores this map at 25 equally spaced scores from 2 below the lowest bin score to 2 above the highest, and scorers interpolate linearly between them. Its risk bands are multiples of the training prevalence $\bar{p} = \sum_k \tilde{y}_k / \sum_k \tilde{m}_k$: Typical up to $1.5\bar{p}$, Elevated up to $3\bar{p}$, and High above.

*Elastic net.* Inside the enclave, we standardized every candidate feature, eligible or not, with its training mean and standard deviation, $z_{ij} = (x_{ij} - \bar{x}_j)/\text{sd}_j$, and solved

$$\min_{w_0,\, w} \; \frac{1-\rho}{2}\|w\|_2^2 + \rho\|w\|_1 + C\sum_i \log\Big[1 + \exp\{-\ell_i(w_0 + z_i^{\mathsf{T}} w)\}\Big],$$

with $\ell_i = 2L_i - 1$, mixing parameter $\rho = 0.5$, inverse penalty strength $C = 0.05$, and an unpenalized intercept (scikit-learn LogisticRegression with the SAGA solver and at most 5,000 iterations), with no tuning and no class weights. Its score is the fitted probability $\sigma(w_0 + z^{\mathsf{T}} w)$.

*Evaluation measures.* In the validation split, with scores $s_i$ and a target $T_i \in \{0, 1\}$ (self-report, uncoded self-report, or a later code) with $N_1$ positives and $N_0$ negatives, the AUROC is the probability that a random positive scores above a random negative, with ties counted as one half:

$$\text{AUROC} = \frac{1}{N_1 N_0} \sum_{i:T_i=1} \; \sum_{k:T_k=0} \Big[\mathbf{1}(s_i > s_k) + \tfrac{1}{2}\,\mathbf{1}(s_i = s_k)\Big].$$

Average precision is $\text{AP} = \sum_t (R_t - R_{t-1}) P_t$ over the distinct score thresholds $t$, where $R_t$ and $P_t$ are the recall and precision of flagging every participant at or above $t$. It equals the target prevalence for a score unrelated to the target. Recall in the top fraction $f \in \{0.05, 0.10\}$ ranks all $n_v$ validation participants by score, takes the first $K = \text{round}(f\, n_v)$, and divides the number of positives among them by $N_1$. On the main split, used for Table 1 of the main text, ties at the cutoff were broken by sort order, and on the second split by a hash of the participant identifier. Intervals are the 2.5th and 97.5th percentiles over $B = 200$ participant-level bootstrap resamples of the validation split with the fitted models held fixed, skipping resamples with one class missing. A paired difference $\Delta = M(s^A) - M(s^B)$ between scores $A$ and $B$ was computed on each resample, so both scores saw the same participants. For recall, the paired comparisons counted participants at or above the $(1 - f)$ quantile of each score in the resample.

*Sensitivity analyses.* A second 80/20 split assigned participants by a salted SHA-256 hash of their identifier, so that it reproduces exactly. We refit all four models on its training part, with a fixed random state for the elastic-net solver and a fixed bootstrap seed (Table S6). We then changed one element at a time. (1) Target: self-report among validation participants with no later code, so that later-coded self-reporters are removed instead of counted as negatives. (2) Self-report timing: the label limited to a first-told answer dated by day 30, with later-only reporters counted as non-reporters and the self-report score refit. (3) Follow-up: validation participants with at least 730 days from day 0 to their last recorded visit, with a code within 730 days and self-report without a code within 730 days as targets. (4) Features and counts: both summary scores fitted from rounded counts of the features eligible for both labels, 69 for chronic fatigue, 73 for fibromyalgia, and 77 for endometriosis. (5) Baselines: a logistic regression on age, sex, and the logarithm of 1 plus the number of visits in the year and in the 5 years before day 0, trained on each label, and the number of symptom groups coded, which uses no label. Subgroups were sex, age (18–44, 45–64, and 65 years or older), and whether day 0 fell before July 1, 2021 (Table S7). For endometriosis, we also repeated the analysis among women aged 18 to 44 years. Table S5 gives the participant flow.

*Calibration of the exported scores.* We applied each exported score and its calibration map, fitted as above on the training part of the main split, to its validation part and fitted two logistic models for self-report by Newton–Raphson with Wald intervals: the calibration intercept $\alpha$ in $\operatorname{logit} P(L = 1 \mid x) = \alpha + \operatorname{logit} \hat{p}(x)$, with the map's linear predictor as an offset, and the calibration slope $\gamma$ in $\operatorname{logit} P(L = 1 \mid x) = \alpha' + \gamma \operatorname{logit} \hat{p}(x)$. The intercepts were −0.03 (95% CI, −0.08 to 0.03) for chronic fatigue, −0.13 (−0.20 to −0.07) for fibromyalgia, and −0.02 (−0.07 to 0.03) for endometriosis, and the slopes were 0.89 (0.85–0.94), 0.84 (0.77–0.90), and 0.98 (0.84–1.11). A negative intercept means the map overestimates the probability of self-report on average, and a slope below 1 means its predictions are too spread out.

**Table S1: Candidate features for the chronic fatigue model.** Registry unm-chronicfatigue-1.0.0 (version 1), 73 candidate binary features. The label codes (G93.3, R53.82) are never features, and participants with a label code on or before day 0 were not scored. Symptom groups: autonomic, cognitive, fatigue, gastrointestinal, immune, mood, pain, sleep. All 73 features met the eligibility rule for the self-report label (main text, Models). Window: 5 y covers days 1,825 to 1 before day 0, and 1 y covers days 365 to 1 before day 0. We evaluated demographics at day 0. ICD-10-CM codes match by prefix on the source code in the condition table and, for the 35 features marked *, also in the observation table. A symptom-group count is 1 when diagnosis-code features from at least the stated number of symptom groups are recorded. CPT and HCPCS codes match source codes in the procedure table exactly (ranges inclusive). A laboratory feature is any recorded measurement of a standard LOINC measurement concept whose name contains the text shown (case-insensitive). A medication feature is any drug exposure record for an RxNorm or RxNorm Extension concept descending from the listed ingredients. Visit classes come from the visit type: inpatient, emergency, or outpatient (all other visit types except pharmacy and laboratory). A feature is 1 if a qualifying record exists in its window and 0 otherwise. A feature entered a model only if it met the eligibility rule in the main text (Models).

| Feature ID | Label | Window | Codes by system |
|---|---|---|---|
| *Demographics* | | | |
| `age_18_39` | Age 18–39 | Day 0 | Age at day 0 (calendar year minus birth year), 18 to 39 years |
| `age_40_49` | Age 40–49 | Day 0 | Age at day 0 (calendar year minus birth year), 40 to 49 years |
| `age_50_59` | Age 50–59 | Day 0 | Age at day 0 (calendar year minus birth year), 50 to 59 years |
| `age_60_69` | Age 60–69 | Day 0 | Age at day 0 (calendar year minus birth year), 60 to 69 years |
| `age_70p` | Age 70 or older | Day 0 | Age at day 0 (calendar year minus birth year), 70 years or older |
| `sex_female` | Female | Day 0 | Sex at birth recorded as female |
| *Diagnosis codes* | | | |
| `cx_fatigue_other`* | Fatigue or malaise coded | 5 y | ICD-10-CM: R53.0, R53.1, R53.81, R53.83<br>Symptom group: fatigue |
| `cx_fatigue_recent`* | Fatigue or malaise coded in the past year | 1 y | ICD-10-CM: R53.0, R53.1, R53.81, R53.83 |
| `cx_sleep`* | Sleep disturbance | 5 y | ICD-10-CM: G47.0, G47.1, G47.8, G47.9, F51<br>Symptom group: sleep |
| `cx_sleep_apnea`* | Sleep apnea | 5 y | ICD-10-CM: G47.33 |
| `cx_cognitive`* | Cognitive symptoms (brain fog, memory) | 5 y | ICD-10-CM: R41, G31.84<br>Symptom group: cognitive |
| `cx_autonomic`* | Palpitations, POTS, orthostatic hypotension, or dysautonomia | 5 y | ICD-10-CM: R00.0, R00.2, I49.8, G90.A, I95.1, G90.9<br>Symptom group: autonomic |
| `cx_dizzy_syncope`* | Dizziness or fainting | 5 y | ICD-10-CM: R42, R55<br>Symptom group: autonomic |
| `cx_headache`* | Headache | 5 y | ICD-10-CM: R51, G44<br>Symptom group: pain |
| `cx_migraine`* | Migraine | 5 y | ICD-10-CM: G43 |
| `cx_myalgia_arthralgia`* | Muscle or joint pain | 5 y | ICD-10-CM: M79.1, M25.5, M79.6<br>Symptom group: pain |
| `cx_fibromyalgia`* | Fibromyalgia | 5 y | ICD-10-CM: M79.7 |
| `cx_chronic_pain`* | Chronic pain | 5 y | ICD-10-CM: G89 |

**Table S1:** (continued)

| Feature ID | Label | Window | Codes by system |
|---|---|---|---|
| cx_depression* | Depression | 5 y | ICD-10-CM: F32, F33<br>Symptom group: mood |
| cx_anxiety* | Anxiety | 5 y | ICD-10-CM: F41<br>Symptom group: mood |
| cx_ptsd* | Posttraumatic stress disorder | 5 y | ICD-10-CM: F43.1 |
| cx_ibs_gi* | Irritable bowel or gastrointestinal symptoms | 5 y | ICD-10-CM: K58, R10, R11, R14, R19<br>Symptom group: gastrointestinal |
| cx_sore_throat_nodes* | Sore throat or enlarged lymph nodes | 5 y | ICD-10-CM: J02, R59<br>Symptom group: immune |
| cx_fever* | Fever | 5 y | ICD-10-CM: R50<br>Symptom group: immune |
| cx_ebv_mono* | Infectious mononucleosis (EBV) | 5 y | ICD-10-CM: B27 |
| cx_lyme* | Lyme disease | 5 y | ICD-10-CM: A69.2 |
| cx_covid* | COVID-19 | 5 y | ICD-10-CM: U07.1 |
| cx_long_covid* | Long COVID | 5 y | ICD-10-CM: U09.9 |
| cx_eds_hypermobility* | Ehlers-Danlos syndrome or hypermobility | 5 y | ICD-10-CM: Q79.6, M35.7 |
| cx_mast_cell* | Mast cell disorder | 5 y | ICD-10-CM: D89.4 |
| cx_hypothyroid* | Hypothyroidism | 5 y | ICD-10-CM: E03 |
| cx_anemia* | Anemia | 5 y | ICD-10-CM: D50, D51, D52, D53, D63, D64 |
| cx_vitd_deficiency* | Vitamin D deficiency | 5 y | ICD-10-CM: E55 |
| cx_autoimmune* | Autoimmune or demyelinating disease | 5 y | ICD-10-CM: M05, M06, M32, M35, L40, K50, K51, M45, G35 |
| cx_obesity* | Obesity | 5 y | ICD-10-CM: E66, Z68.3, Z68.4 |
| cx_diabetes* | Diabetes | 5 y | ICD-10-CM: E10, E11 |
| cx_hypertension* | Hypertension | 5 y | ICD-10-CM: I10 |
| cx_heart_disease* | Coronary disease or heart failure | 5 y | ICD-10-CM: I25, I50 |
| cx_cancer* | Cancer | 5 y | ICD-10-CM: C |
| *Symptom-group counts* | | | |
| clusters_ge3* | Three or more symptom types | 5 y | Number of symptom groups recorded: 3 or more |
| clusters_ge5* | Five or more symptom types | 5 y | Number of symptom groups recorded: 5 or more |
| *Procedures* | | | |
| px_tilt | Tilt-table test | 5 y | CPT or HCPCS: 93660 |
| px_sleep_study | Sleep study | 5 y | CPT or HCPCS: 95806–95811, G0398, G0399, G0400 |
| px_brain_mri | Brain MRI | 5 y | CPT or HCPCS: 70551–70553 |
| px_echo | Echocardiogram | 5 y | CPT or HCPCS: 93303–93308, 93312–93318, 93350–93352 |
| px_cardiac_monitor | Holter or event monitor | 5 y | CPT or HCPCS: 93224–93229, 93241–93248, 93268–93272 |
| px_exercise_test | Exercise or cardiopulmonary exercise test | 5 y | CPT or HCPCS: 94621, 93015–93018 |

**Table S1:** (continued)

| Feature ID | Label | Window | Codes by system |
|---|---|---|---|
| px_neuropsych | Neuropsychological testing | 5 y | CPT or HCPCS: 96132, 96133, 96136–96139 |
| px_pt_eval | Physical therapy evaluation | 5 y | CPT or HCPCS: 97161–97163 |
| *Laboratory tests* | | | |
| lab_tsh | Thyroid (TSH) test | 5 y | LOINC: measurement concepts whose name contains "Thyrotropin" |
| lab_ferritin | Ferritin test | 5 y | LOINC: measurement concepts whose name contains "Ferritin" |
| lab_b12 | Vitamin B12 test | 5 y | LOINC: measurement concepts whose name contains "Cobalamin" |
| lab_vitamin_d | Vitamin D test | 5 y | LOINC: measurement concepts whose name contains "25-hydroxyvitamin D" |
| lab_ana | Antinuclear antibody (ANA) test | 5 y | LOINC: measurement concepts whose name contains "Nuclear Ab" |
| lab_crp | C-reactive protein test | 5 y | LOINC: measurement concepts whose name contains "C reactive protein" |
| lab_esr | Sedimentation rate (ESR) test | 5 y | LOINC: measurement concepts whose name contains "Erythrocyte sedimentation rate" |
| lab_ebv | Epstein-Barr virus test | 5 y | LOINC: measurement concepts whose name contains "Epstein Barr virus" |
| lab_lyme | Lyme disease test | 5 y | LOINC: measurement concepts whose name contains "Borrelia burgdorferi" |
| lab_cortisol | Cortisol test | 5 y | LOINC: measurement concepts whose name contains "Cortisol" |
| lab_ck | Creatine kinase test | 5 y | LOINC: measurement concepts whose name contains "Creatine kinase" |
| *Medications* | | | |
| rx_stimulant | Stimulant or wakefulness drug | 5 y | RxNorm ingredients: modafinil, armodafinil, methylphenidate, amphetamine, dextroamphetamine, lisdexamfetamine |
| rx_naltrexone | Naltrexone | 5 y | RxNorm ingredients: naltrexone |
| rx_ssri_snri | SSRI or SNRI antidepressant | 5 y | RxNorm ingredients: sertraline, fluoxetine, citalopram, escitalopram, paroxetine, venlafaxine, duloxetine, desvenlafaxine |
| rx_tricyclic_lowdose | Amitriptyline, nortriptyline, or cyclobenzaprine | 5 y | RxNorm ingredients: amitriptyline, nortriptyline, cyclobenzaprine |
| rx_gabapentinoid | Gabapentin or pregabalin | 5 y | RxNorm ingredients: gabapentin, pregabalin |
| rx_sleep_aid | Sleep medication | 5 y | RxNorm ingredients: zolpidem, trazodone, eszopiclone |
| rx_beta_blocker | Beta blocker | 5 y | RxNorm ingredients: propranolol, metoprolol, atenolol, bisoprolol, nadolol, carvedilol |
| rx_pots | POTS medication | 5 y | RxNorm ingredients: ivabradine, midodrine, fludrocortisone, pyridostigmine |
| rx_opioid | Opioid | 5 y | RxNorm ingredients: oxycodone, hydrocodone, tramadol, morphine |

**Table S1:** (continued)

| Feature ID | Label | Window | Codes by system |
|---|---|---|---|
| *Health care use* | | | |
| `util_out_r_ge6` | 6+ outpatient visits in the past year | 1 y | Outpatient visits: 6 or more |
| `util_out_r_ge12` | 12+ outpatient visits in the past year | 1 y | Outpatient visits: 12 or more |
| `util_ed_r` | Emergency visit in the past year | 1 y | Emergency visits: 1 or more |
| `util_ip_h` | Hospital stay in the past 5 years | 5 y | Inpatient visits: 1 or more |

**Table S2: Candidate features for the fibromyalgia model.** Registry unm-fibromyalgia-1.0.0 (version 1), 83 candidate binary features. The label codes (M79.7) are never features, and participants with a label code on or before day 0 were not scored. Symptom groups: autonomic, cognitive, fatigue, gastrointestinal, immune, mood, pain, sleep. Of the 83 features, 80 met the eligibility rule for the self-report label (main text, Models). The exceptions were `cx_ebv_mono`, `cx_mast_cell`, and `px_tilt`. Windows, matching rules, and eligibility as in Table S1, except that all diagnosis-code and symptom-group features use both the condition and observation tables.

| Feature ID | Label | Window | Codes by system |
|---|---|---|---|
| *Demographics* | | | |
| `age_18_39` | Age 18–39 | Day 0 | Age at day 0 (calendar year minus birth year), 18 to 39 years |
| `age_40_49` | Age 40–49 | Day 0 | Age at day 0 (calendar year minus birth year), 40 to 49 years |
| `age_50_59` | Age 50–59 | Day 0 | Age at day 0 (calendar year minus birth year), 50 to 59 years |
| `age_60_69` | Age 60–69 | Day 0 | Age at day 0 (calendar year minus birth year), 60 to 69 years |
| `age_70p` | Age 70 or older | Day 0 | Age at day 0 (calendar year minus birth year), 70 years or older |
| `sex_female` | Female | Day 0 | Sex at birth recorded as female |
| *Diagnosis codes* | | | |
| `cx_fatigue_other` | Fatigue or malaise coded | 5 y | ICD-10-CM: R53.0, R53.1, R53.81, R53.83<br>Symptom group: fatigue |
| `cx_fatigue_recent` | Fatigue or malaise coded in the past year | 1 y | ICD-10-CM: R53.0, R53.1, R53.81, R53.83 |
| `cx_sleep` | Sleep disturbance | 5 y | ICD-10-CM: G47.0, G47.1, G47.8, G47.9, F51<br>Symptom group: sleep |
| `cx_sleep_apnea` | Sleep apnea | 5 y | ICD-10-CM: G47.33 |
| `cx_cognitive` | Cognitive symptoms (brain fog, memory) | 5 y | ICD-10-CM: R41, G31.84<br>Symptom group: cognitive |
| `cx_autonomic` | Palpitations, POTS, orthostatic hypotension, or dysautonomia | 5 y | ICD-10-CM: R00.0, R00.2, I49.8, G90.A, I95.1, G90.9<br>Symptom group: autonomic |
| `cx_dizzy_syncope` | Dizziness or fainting | 5 y | ICD-10-CM: R42, R55<br>Symptom group: autonomic |
| `cx_headache` | Headache | 5 y | ICD-10-CM: R51, G44<br>Symptom group: pain |
| `cx_migraine` | Migraine | 5 y | ICD-10-CM: G43 |
| `cx_myalgia_arthralgia` | Muscle or joint pain | 5 y | ICD-10-CM: M79.1, M25.5, M79.6<br>Symptom group: pain |
| `cx_chronic_pain` | Chronic pain | 5 y | ICD-10-CM: G89 |
| `cx_depression` | Depression | 5 y | ICD-10-CM: F32, F33<br>Symptom group: mood |
| `cx_anxiety` | Anxiety | 5 y | ICD-10-CM: F41<br>Symptom group: mood |
| `cx_ptsd` | Posttraumatic stress disorder | 5 y | ICD-10-CM: F43.1 |
| `cx_ibs_gi` | Irritable bowel or gastrointestinal symptoms | 5 y | ICD-10-CM: K58, R10, R11, R14, R19<br>Symptom group: gastrointestinal |
| `cx_sore_throat_nodes` | Sore throat or enlarged lymph nodes | 5 y | ICD-10-CM: J02, R59<br>Symptom group: immune |

**Table S2:** (continued)

| Feature ID | Label | Window | Codes by system |
|---|---|---|---|
| cx_fever | Fever | 5 y | ICD-10-CM: R50<br>Symptom group: immune |
| cx_ebv_mono | Infectious mononucleosis (EBV) | 5 y | ICD-10-CM: B27 |
| cx_lyme | Lyme disease | 5 y | ICD-10-CM: A69.2 |
| cx_covid | COVID-19 | 5 y | ICD-10-CM: U07.1 |
| cx_long_covid | Long COVID | 5 y | ICD-10-CM: U09.9 |
| cx_eds_hypermobility | Ehlers-Danlos syndrome or hypermobility | 5 y | ICD-10-CM: Q79.6, M35.7 |
| cx_mast_cell | Mast cell disorder | 5 y | ICD-10-CM: D89.4 |
| cx_hypothyroid | Hypothyroidism | 5 y | ICD-10-CM: E03 |
| cx_anemia | Anemia | 5 y | ICD-10-CM: D50, D51, D52, D53, D63, D64 |
| cx_vitd_deficiency | Vitamin D deficiency | 5 y | ICD-10-CM: E55 |
| cx_autoimmune | Autoimmune or demyelinating disease | 5 y | ICD-10-CM: M05, M06, M32, M35, L40, K50, K51, M45, G35 |
| cx_obesity | Obesity | 5 y | ICD-10-CM: E66, Z68.3, Z68.4 |
| cx_diabetes | Diabetes | 5 y | ICD-10-CM: E10, E11 |
| cx_hypertension | Hypertension | 5 y | ICD-10-CM: I10 |
| cx_heart_disease | Coronary disease or heart failure | 5 y | ICD-10-CM: I25, I50 |
| cx_cancer | Cancer | 5 y | ICD-10-CM: C |
| cx_cfs | Chronic fatigue or ME/CFS | 5 y | ICD-10-CM: G93.3, R53.82<br>Symptom group: fatigue |
| cx_tmj | Temporomandibular joint disorder | 5 y | ICD-10-CM: M26.6<br>Symptom group: pain |
| cx_rls | Restless legs syndrome | 5 y | ICD-10-CM: G25.81<br>Symptom group: sleep |
| cx_interstitial_cystitis | Interstitial cystitis or bladder pain | 5 y | ICD-10-CM: N30.1 |
| cx_osteoarthritis | Osteoarthritis | 5 y | ICD-10-CM: M15, M16, M17, M18, M19 |
| cx_neck_back_pain | Neck or low back pain | 5 y | ICD-10-CM: M54.2, M54.5<br>Symptom group: pain |
| *Symptom-group counts* | | | |
| clusters_ge3 | Three or more symptom types | 5 y | Number of symptom groups recorded: 3 or more |
| clusters_ge5 | Five or more symptom types | 5 y | Number of symptom groups recorded: 5 or more |
| *Procedures* | | | |
| px_tilt | Tilt-table test | 5 y | CPT or HCPCS: 93660 |
| px_sleep_study | Sleep study | 5 y | CPT or HCPCS: 95806–95811, G0398, G0399, G0400 |
| px_brain_mri | Brain MRI | 5 y | CPT or HCPCS: 70551–70553 |
| px_echo | Echocardiogram | 5 y | CPT or HCPCS: 93303–93308, 93312–93318, 93350–93352 |
| px_cardiac_monitor | Holter or event monitor | 5 y | CPT or HCPCS: 93224–93229, 93241–93248, 93268–93272 |
| px_exercise_test | Exercise or cardiopulmonary exercise test | 5 y | CPT or HCPCS: 94621, 93015–93018 |

**Table S2:** (continued)

| Feature ID | Label | Window | Codes by system |
|---|---|---|---|
| px_neuropsych | Neuropsychological testing | 5 y | CPT or HCPCS: 96132, 96133, 96136–96139 |
| px_pt_eval | Physical therapy evaluation | 5 y | CPT or HCPCS: 97161–97163 |
| *Laboratory tests* | | | |
| lab_tsh | Thyroid (TSH) test | 5 y | LOINC: measurement concepts whose name contains "Thyrotropin" |
| lab_ferritin | Ferritin test | 5 y | LOINC: measurement concepts whose name contains "Ferritin" |
| lab_b12 | Vitamin B12 test | 5 y | LOINC: measurement concepts whose name contains "Cobalamin" |
| lab_vitamin_d | Vitamin D test | 5 y | LOINC: measurement concepts whose name contains "25-hydroxyvitamin D" |
| lab_ana | Antinuclear antibody (ANA) test | 5 y | LOINC: measurement concepts whose name contains "Nuclear Ab" |
| lab_crp | C-reactive protein test | 5 y | LOINC: measurement concepts whose name contains "C reactive protein" |
| lab_esr | Sedimentation rate (ESR) test | 5 y | LOINC: measurement concepts whose name contains "Erythrocyte sedimentation rate" |
| lab_ebv | Epstein-Barr virus test | 5 y | LOINC: measurement concepts whose name contains "Epstein Barr virus" |
| lab_lyme | Lyme disease test | 5 y | LOINC: measurement concepts whose name contains "Borrelia burgdorferi" |
| lab_cortisol | Cortisol test | 5 y | LOINC: measurement concepts whose name contains "Cortisol" |
| lab_ck | Creatine kinase test | 5 y | LOINC: measurement concepts whose name contains "Creatine kinase" |
| lab_rf | Rheumatoid factor test | 5 y | LOINC: measurement concepts whose name contains "Rheumatoid factor" |
| lab_ccp | Anti-CCP antibody test | 5 y | LOINC: measurement concepts whose name contains "Cyclic citrullinated peptide" |
| *Medications* | | | |
| rx_stimulant | Stimulant or wakefulness drug | 5 y | RxNorm ingredients: modafinil, armodafinil, methylphenidate, amphetamine, dextroamphetamine, lisdexamfetamine |
| rx_naltrexone | Naltrexone | 5 y | RxNorm ingredients: naltrexone |
| rx_ssri_snri | SSRI or SNRI antidepressant | 5 y | RxNorm ingredients: sertraline, fluoxetine, citalopram, escitalopram, paroxetine, venlafaxine, duloxetine, desvenlafaxine |
| rx_tricyclic_lowdose | Amitriptyline, nortriptyline, or cyclobenzaprine | 5 y | RxNorm ingredients: amitriptyline, nortriptyline, cyclobenzaprine |
| rx_gabapentinoid | Gabapentin or pregabalin | 5 y | RxNorm ingredients: gabapentin, pregabalin |
| rx_sleep_aid | Sleep medication | 5 y | RxNorm ingredients: zolpidem, trazodone, eszopiclone |
| rx_beta_blocker | Beta blocker | 5 y | RxNorm ingredients: propranolol, metoprolol, atenolol, bisoprolol, nadolol, carvedilol |

**Table S2:** (continued)

| Feature ID | Label | Window | Codes by system |
|---|---|---|---|
| rx_pots | POTS medication | 5 y | RxNorm ingredients: ivabradine, midodrine, fludrocortisone, pyridostigmine |
| rx_opioid | Opioid | 5 y | RxNorm ingredients: oxycodone, hydrocodone, tramadol, morphine |
| rx_milnacipran | Milnacipran | 5 y | RxNorm ingredients: milnacipran |
| rx_nsaid | NSAID pain reliever | 5 y | RxNorm ingredients: naproxen, ibuprofen, meloxicam, diclofenac, ketorolac |
| rx_muscle_relaxant | Muscle relaxant | 5 y | RxNorm ingredients: tizanidine, methocarbamol, baclofen |
| *Health care use* | | | |
| util_out_r_ge6 | 6+ outpatient visits in the past year | 1 y | Outpatient visits: 6 or more |
| util_out_r_ge12 | 12+ outpatient visits in the past year | 1 y | Outpatient visits: 12 or more |
| util_ed_r | Emergency visit in the past year | 1 y | Emergency visits: 1 or more |
| util_ip_h | Hospital stay in the past 5 years | 5 y | Inpatient visits: 1 or more |

**Table S3: Candidate features for the endometriosis model.** Registry unm-endometriosis-1.0.0 (version 1), 94 candidate binary features. The label codes (N80) are never features, and participants with a label code on or before day 0 were not scored. Symptom groups: autonomic, cognitive, fatigue, gastrointestinal, immune, menstrual, mood, pain, sleep, urinary. Of the 94 features, 90 met the eligibility rule for the self-report label (main text, Models). The exceptions were `sex_female`, `cx_ebv_mono`, `px_diag_laparoscopy`, and `px_lap_excision`. Windows, matching rules, and eligibility as in Table S1, except that all diagnosis-code and symptom-group features use both the condition and observation tables.

| Feature ID | Label | Window | Codes by system |
|---|---|---|---|
| *Demographics* | | | |
| `age_18_39` | Age 18–39 | Day 0 | Age at day 0 (calendar year minus birth year), 18 to 39 years |
| `age_40_49` | Age 40–49 | Day 0 | Age at day 0 (calendar year minus birth year), 40 to 49 years |
| `age_50_59` | Age 50–59 | Day 0 | Age at day 0 (calendar year minus birth year), 50 to 59 years |
| `age_60_69` | Age 60–69 | Day 0 | Age at day 0 (calendar year minus birth year), 60 to 69 years |
| `age_70p` | Age 70 or older | Day 0 | Age at day 0 (calendar year minus birth year), 70 years or older |
| `sex_female` | Female | Day 0 | Sex at birth recorded as female |
| *Diagnosis codes* | | | |
| `cx_fatigue_other` | Fatigue or malaise coded | 5 y | ICD-10-CM: R53.0, R53.1, R53.81, R53.83<br>Symptom group: fatigue |
| `cx_fatigue_recent` | Fatigue or malaise coded in the past year | 1 y | ICD-10-CM: R53.0, R53.1, R53.81, R53.83 |
| `cx_sleep` | Sleep disturbance | 5 y | ICD-10-CM: G47.0, G47.1, G47.8, G47.9, F51<br>Symptom group: sleep |
| `cx_sleep_apnea` | Sleep apnea | 5 y | ICD-10-CM: G47.33 |
| `cx_cognitive` | Cognitive symptoms (brain fog, memory) | 5 y | ICD-10-CM: R41, G31.84<br>Symptom group: cognitive |
| `cx_autonomic` | Palpitations, POTS, orthostatic hypotension, or dysautonomia | 5 y | ICD-10-CM: R00.0, R00.2, I49.8, G90.A, I95.1, G90.9<br>Symptom group: autonomic |
| `cx_dizzy_syncope` | Dizziness or fainting | 5 y | ICD-10-CM: R42, R55<br>Symptom group: autonomic |
| `cx_headache` | Headache | 5 y | ICD-10-CM: R51, G44<br>Symptom group: pain |
| `cx_migraine` | Migraine | 5 y | ICD-10-CM: G43 |
| `cx_myalgia_arthralgia` | Muscle or joint pain | 5 y | ICD-10-CM: M79.1, M25.5, M79.6<br>Symptom group: pain |
| `cx_fibromyalgia` | Fibromyalgia | 5 y | ICD-10-CM: M79.7 |
| `cx_chronic_pain` | Chronic pain | 5 y | ICD-10-CM: G89 |
| `cx_depression` | Depression | 5 y | ICD-10-CM: F32, F33<br>Symptom group: mood |
| `cx_anxiety` | Anxiety | 5 y | ICD-10-CM: F41<br>Symptom group: mood |
| `cx_ptsd` | Posttraumatic stress disorder | 5 y | ICD-10-CM: F43.1 |
| `cx_ibs_gi` | Irritable bowel or gastrointestinal symptoms | 5 y | ICD-10-CM: K58, R10, R11, R14, R19<br>Symptom group: gastrointestinal |

**Table S3:** (continued)

| Feature ID | Label | Window | Codes by system |
|---|---|---|---|
| cx_sore_throat_nodes | Sore throat or enlarged lymph nodes | 5 y | ICD-10-CM: J02, R59<br>Symptom group: immune |
| cx_fever | Fever | 5 y | ICD-10-CM: R50<br>Symptom group: immune |
| cx_ebv_mono | Infectious mononucleosis (EBV) | 5 y | ICD-10-CM: B27 |
| cx_lyme | Lyme disease | 5 y | ICD-10-CM: A69.2 |
| cx_covid | COVID-19 | 5 y | ICD-10-CM: U07.1 |
| cx_long_covid | Long COVID | 5 y | ICD-10-CM: U09.9 |
| cx_eds_hypermobility | Ehlers-Danlos syndrome or hypermobility | 5 y | ICD-10-CM: Q79.6, M35.7 |
| cx_mast_cell | Mast cell disorder | 5 y | ICD-10-CM: D89.4 |
| cx_hypothyroid | Hypothyroidism | 5 y | ICD-10-CM: E03 |
| cx_anemia | Anemia | 5 y | ICD-10-CM: D50, D51, D52, D53, D63, D64 |
| cx_vitd_deficiency | Vitamin D deficiency | 5 y | ICD-10-CM: E55 |
| cx_autoimmune | Autoimmune or demyelinating disease | 5 y | ICD-10-CM: M05, M06, M32, M35, L40, K50, K51, M45, G35 |
| cx_obesity | Obesity | 5 y | ICD-10-CM: E66, Z68.3, Z68.4 |
| cx_diabetes | Diabetes | 5 y | ICD-10-CM: E10, E11 |
| cx_hypertension | Hypertension | 5 y | ICD-10-CM: I10 |
| cx_heart_disease | Coronary disease or heart failure | 5 y | ICD-10-CM: I25, I50 |
| cx_cancer | Cancer | 5 y | ICD-10-CM: C |
| cx_pelvic_pain | Pelvic pain | 5 y | ICD-10-CM: R10.2<br>Symptom group: pain |
| cx_dysmenorrhea | Painful periods (dysmenorrhea) | 5 y | ICD-10-CM: N94.4, N94.5, N94.6<br>Symptom group: menstrual |
| cx_dyspareunia | Pain with intercourse | 5 y | ICD-10-CM: N94.1<br>Symptom group: pain |
| cx_menstrual | Heavy or irregular menstrual bleeding | 5 y | ICD-10-CM: N92<br>Symptom group: menstrual |
| cx_infertility | Female infertility | 5 y | ICD-10-CM: N97 |
| cx_ovarian_cyst | Ovarian cyst | 5 y | ICD-10-CM: N83.2 |
| cx_fibroids | Uterine fibroids | 5 y | ICD-10-CM: D25 |
| cx_pcos | Polycystic ovary syndrome | 5 y | ICD-10-CM: E28.2 |
| cx_interstitial_cystitis | Interstitial cystitis or bladder pain | 5 y | ICD-10-CM: N30.1<br>Symptom group: urinary |
| cx_back_pain | Low back pain | 5 y | ICD-10-CM: M54.5<br>Symptom group: pain |
| cx_constipation | Constipation | 5 y | ICD-10-CM: K59.0<br>Symptom group: gastrointestinal |
| *Symptom-group counts* | | | |
| clusters_ge3 | Three or more symptom types | 5 y | Number of symptom groups recorded: 3 or more |
| clusters_ge5 | Five or more symptom types | 5 y | Number of symptom groups recorded: 5 or more |
| *Procedures* | | | |
| px_tilt | Tilt-table test | 5 y | CPT or HCPCS: 93660 |

**Table S3:** (continued)

| Feature ID | Label | Window | Codes by system |
|---|---|---|---|
| px_sleep_study | Sleep study | 5 y | CPT or HCPCS: 95806–95811, G0398, G0399, G0400 |
| px_brain_mri | Brain MRI | 5 y | CPT or HCPCS: 70551–70553 |
| px_echo | Echocardiogram | 5 y | CPT or HCPCS: 93303–93308, 93312–93318, 93350–93352 |
| px_cardiac_monitor | Holter or event monitor | 5 y | CPT or HCPCS: 93224–93229, 93241–93248, 93268–93272 |
| px_exercise_test | Exercise or cardiopulmonary exercise test | 5 y | CPT or HCPCS: 94621, 93015–93018 |
| px_neuropsych | Neuropsychological testing | 5 y | CPT or HCPCS: 96132, 96133, 96136–96139 |
| px_pt_eval | Physical therapy evaluation | 5 y | CPT or HCPCS: 97161–97163 |
| px_pelvic_us | Pelvic or transvaginal ultrasound | 5 y | CPT or HCPCS: 76830, 76856, 76857 |
| px_diag_laparoscopy | Diagnostic laparoscopy | 5 y | CPT or HCPCS: 49320 |
| px_lap_excision | Laparoscopic excision or fulguration of pelvic lesions | 5 y | CPT or HCPCS: 58662 |
| px_hysterectomy | Hysterectomy | 5 y | CPT or HCPCS: 58150, 58570–58573 |
| px_pelvic_mri | Pelvic MRI | 5 y | CPT or HCPCS: 72195–72197 |
| *Laboratory tests* | | | |
| lab_tsh | Thyroid (TSH) test | 5 y | LOINC: measurement concepts whose name contains “Thyrotropin” |
| lab_ferritin | Ferritin test | 5 y | LOINC: measurement concepts whose name contains “Ferritin” |
| lab_b12 | Vitamin B12 test | 5 y | LOINC: measurement concepts whose name contains “Cobalamin” |
| lab_vitamin_d | Vitamin D test | 5 y | LOINC: measurement concepts whose name contains “25-hydroxyvitamin D” |
| lab_ana | Antinuclear antibody (ANA) test | 5 y | LOINC: measurement concepts whose name contains “Nuclear Ab” |
| lab_crp | C-reactive protein test | 5 y | LOINC: measurement concepts whose name contains “C reactive protein” |
| lab_esr | Sedimentation rate (ESR) test | 5 y | LOINC: measurement concepts whose name contains “Erythrocyte sedimentation rate” |
| lab_ebv | Epstein-Barr virus test | 5 y | LOINC: measurement concepts whose name contains “Epstein Barr virus” |
| lab_lyme | Lyme disease test | 5 y | LOINC: measurement concepts whose name contains “Borrelia burgdorferi” |
| lab_cortisol | Cortisol test | 5 y | LOINC: measurement concepts whose name contains “Cortisol” |
| lab_ck | Creatine kinase test | 5 y | LOINC: measurement concepts whose name contains “Creatine kinase” |
| lab_ca125 | CA-125 test | 5 y | LOINC: measurement concepts whose name contains “Cancer Ag 125” |
| *Medications* | | | |

**Table S3:** (continued)

| Feature ID | Label | Window | Codes by system |
|---|---|---|---|
| rx_stimulant | Stimulant or wakefulness drug | 5 y | RxNorm ingredients: modafinil, armodafinil, methylphenidate, amphetamine, dextroamphetamine, lisdexamfetamine |
| rx_naltrexone | Naltrexone | 5 y | RxNorm ingredients: naltrexone |
| rx_ssri_snri | SSRI or SNRI antidepressant | 5 y | RxNorm ingredients: sertraline, fluoxetine, citalopram, escitalopram, paroxetine, venlafaxine, duloxetine, desvenlafaxine |
| rx_tricyclic_lowdose | Amitriptyline, nortriptyline, or cyclobenzaprine | 5 y | RxNorm ingredients: amitriptyline, nortriptyline, cyclobenzaprine |
| rx_gabapentinoid | Gabapentin or pregabalin | 5 y | RxNorm ingredients: gabapentin, pregabalin |
| rx_sleep_aid | Sleep medication | 5 y | RxNorm ingredients: zolpidem, trazodone, eszopiclone |
| rx_beta_blocker | Beta blocker | 5 y | RxNorm ingredients: propranolol, metoprolol, atenolol, bisoprolol, nadolol, carvedilol |
| rx_pots | POTS medication | 5 y | RxNorm ingredients: ivabradine, midodrine, fludrocortisone, pyridostigmine |
| rx_opioid | Opioid | 5 y | RxNorm ingredients: oxycodone, hydrocodone, tramadol, morphine |
| rx_hormonal | Hormonal contraceptive or progestin | 5 y | RxNorm ingredients: norethindrone, levonorgestrel, drospirenone, medroxyprogesterone, etonogestrel, desogestrel, norgestimate |
| rx_gnrh | GnRH agonist or antagonist | 5 y | RxNorm ingredients: leuprolide, elagolix, relugolix |
| rx_nsaid | NSAID pain reliever | 5 y | RxNorm ingredients: naproxen, ibuprofen, meloxicam, diclofenac, ketorolac |
| rx_aromatase | Aromatase inhibitor | 5 y | RxNorm ingredients: letrozole, anastrozole |
| *Health care use* | | | |
| util_out_r_ge6 | 6+ outpatient visits in the past year | 1 y | Outpatient visits: 6 or more |
| util_out_r_ge12 | 12+ outpatient visits in the past year | 1 y | Outpatient visits: 12 or more |
| util_ed_r | Emergency visit in the past year | 1 y | Emergency visits: 1 or more |
| util_ip_h | Hospital stay in the past 5 years | 5 y | Inpatient visits: 1 or more |

**Table S4: TRIPOD+AI checklist.** Items of the TRIPOD+AI reporting guideline (Collins et al., BMJ 2024;385:e078378), paraphrased, with the place in the manuscript where we report each. Methods refers to the Materials and Methods section, and section and subsection names are those of the main text. Not reported: the manuscript does not address the item. Notes in parentheses identify partial reporting.

| Item | Topic | Checklist item (paraphrased) | Where reported |
|---|---|---|---|
| *Title and abstract* | | | |
| 1 | Title | Identify the study as developing or evaluating a multivariable prediction model, with the target population and the outcome | Title (names the conditions, training labels, and cohort but does not use the term prediction model) |
| 2 | Abstract | Follow the TRIPOD+AI for Abstracts checklist | Abstract (calibration and registration not reported) |
| *Introduction* | | | |
| 3a | Background | Health care context (diagnostic or prognostic), rationale, and existing models | Introduction, paragraphs 1–3 |
| 3b | Background | Target population, intended purpose in the care pathway, and intended users | Introduction (final paragraph), Statement of significance, and Discussion (paragraph 4) |
| 3c | Background | Known health inequalities between sociodemographic groups | Not reported |
| 4 | Objectives | Objectives, including whether the study develops or evaluates a model, or both | Introduction, final paragraph |
| *Methods* | | | |
| 5a | Data | Data sources for development and evaluation, rationale, and representativeness | Methods: Data source and population |
| 5b | Data | Dates of data collection, start and end of accrual, and end of follow-up | Methods: Data source and population (EHR visit periods and data cutoff), Labels and evaluation targets (follow-up to the data cutoff) |
| 6a | Participants | Study setting, and number and location of centers | Methods: Data source and population (number of contributing health systems not stated) |
| 6b | Participants | Eligibility criteria | Methods: Data source and population |
| 6c | Participants | Treatments received and how they were handled | Methods: Features (medication exposures are predictors) |
| 7 | Data preparation | Preprocessing and quality checks, and whether they were similar across sociodemographic groups | Methods: Features and Models (rounding adjustment), with checks by sociodemographic group not reported |
| 8a | Outcome | Outcome and time horizon, how and when assessed, rationale, and consistency across sociodemographic groups | Methods: Labels and evaluation targets, and Supplementary Methods (730-day horizon and survey timing, Table S6) |
| 8b | Outcome | Qualifications and demographics of outcome assessors, if assessment is subjective | Not reported (labels are survey answers and recorded codes) |
| 8c | Outcome | Blinding of outcome assessment | Not reported |

**Table S4:** (continued)

| Item | Topic | Checklist item (paraphrased) | Where reported |
|---|---|---|---|
| 9a | Predictors | Choice of initial predictors and any preselection before model building | Methods: Features, Models (eligibility rule and number of eligible features), and Tables S1–S3 |
| 9b | Predictors | Definition of all predictors, including how and when they were measured | Methods: Features and Tables S1–S3 |
| 9c | Predictors | Qualifications and demographics of predictor assessors, if measurement is subjective | Not reported (predictors are recorded EHR events) |
| 10 | Sample size | How the study size was arrived at, and whether it was sufficient | Not reported |
| 11 | Missing data | Handling of missing data and reasons for omitting data | Methods: Labels and evaluation targets (skipped answer counted as a report and no answer treated as not reporting), Features (absent record coded 0), and Table S5 (exclusions) |
| 12a | Analytical methods | Use and partitioning of the data for development and evaluation | Methods: Models (80% training split), Evaluation (20% validation split), and Supplementary Methods (a second split defined by a hash of the participant identifier) |
| 12b | Analytical methods | Handling of predictors (functional form, rescaling, transformation, standardization) | Methods: Features (binary indicators) and Models (standardized features for the elastic net) |
| 12c | Analytical methods | Model type and rationale, model-building steps, hyperparameter tuning, and internal validation | Methods: Models and Evaluation |
| 12d | Analytical methods | Handling and quantification of heterogeneity across clusters | Not reported (random split of one cohort) |
| 12e | Analytical methods | Performance measures and plots, with rationale, and comparison of models | Methods: Evaluation, Supplementary Methods, and Table S6 (common feature set and two simple baselines) |
| 12f | Analytical methods | Model updating arising from the evaluation | Not reported |
| 12g | Analytical methods | How predictions were calculated for the evaluation | Methods: Models (score formula) and Supplementary Methods |
| 13 | Class imbalance | Class imbalance methods, rationale, and any recalibration | Methods: Models (no class weighting) |
| 14 | Fairness | Approaches used to address model fairness, and rationale | Not reported |
| 15 | Model output | Model output, and how any classification thresholds were chosen | Methods: Models (mapping to probabilities, Supplementary Methods) and Evaluation (top 5% and 10% of scores) |
| 16 | Training vs evaluation | Differences between development and evaluation data in setting, eligibility, outcome, and predictors | Methods: Models, Evaluation (random split of one cohort), and Limitations |
| 17 | Ethical approval | Ethics committee, and informed consent or its waiver | Methods: Disclosure control and ethics (program-level consent and no project-specific review) |
| *Open science* | | | |

**Table S4:** (continued)

| Item | Topic | Checklist item (paraphrased) | Where reported |
|---|---|---|---|
| 18a | Funding | Source of funding and role of the funders | Funding (no specific funding) |
| 18b | Conflicts of interest | Conflicts of interest and financial disclosures for all authors | Declaration of competing interest |
| 18c | Protocol | Where the protocol can be accessed, or that none was prepared | Not reported |
| 18d | Registration | Registration details, or that the study was not registered | Not reported |
| 18e | Data sharing | Availability of the study data | Data and code availability |
| 18f | Code sharing | Availability of the analysis code | Data and code availability (public repository with the analysis code, aggregate statistics, and the exported models with synthetic test patients) |
| *Patient and public involvement* | | | |
| 19 | Patient and public involvement | Involvement in design, conduct, reporting, interpretation, or dissemination, or a statement of none | Not reported |
| *Results* | | | |
| 20a | Participants | Flow of participants, numbers with and without the outcome, and follow-up time | Results: Cohorts (numbers with and without each label), Results: Sensitivity analyses (follow-up time), and Table S5 (exclusions at each eligibility step) |
| 20b | Participants | Characteristics overall and by setting, including key predictors, outcome events, and missing data | Results: Cohorts and Features associated with self-report (selected characteristics, no characteristics table) |
| 20c | Participants | Comparison of predictor and outcome distributions between development and evaluation data | Not reported |
| 21 | Model development | Numbers of participants and outcome events in each analysis | Results: Cohorts and Table 1 |
| 22 | Model specification | Full model, to allow predictions in new individuals and third-party evaluation, with any access restrictions | Methods: Models (score formula), Supplementary Methods, Data and code availability (exported model files), with weights not tabulated |
| 23a | Model performance | Performance with confidence intervals, including key subgroups | Results: Discrimination by training label and Table 1, Results: Sensitivity analyses, and Tables S6 and S7 (subgroups by sex, age, and day 0 period, with intervals) |
| 23b | Model performance | Heterogeneity of performance across clusters, if examined | Not examined (random split of one cohort, Limitations) |
| 24 | Model updating | Results of any model updating | Not reported |
| *Discussion* | | | |
| 25 | Interpretation | Overall interpretation, including fairness, in light of the objectives and previous studies | Discussion, paragraphs 1–3 (fairness not discussed) |
| 26 | Limitations | Limitations and their effects on bias, uncertainty, and generalizability | Discussion: Limitations |

**Table S4:** (continued)

| Item | Topic | Checklist item (paraphrased) | Where reported |
|---|---|---|---|
| 27a | Usability | How poor-quality or unavailable input data should be assessed and handled in implementation | Methods: Features (absent records coded 0) and Limitations (mapping to FHIR not validated) |
| 27b | Usability | Whether users interact with the input data or the model, and the expertise required | Discussion, paragraph 4 (expertise not stated) |
| 27c | Usability | Next steps for research on applicability and generalizability | Discussion, paragraphs 3–4, and Conclusion |

**Table S5: Participant flow.** Participants remaining after each eligibility step, counted once per participant and rounded up to the next multiple of 10. The analysis cohorts are the participants of Table 1 in the main text. Its totals are sums of the separately rounded training and validation counts, so they can differ from this table by 10. Dates are shifted by 1 to 365 days per participant, so the day 0 period is approximate. –: not applicable.

| Step | Chronic fatigue | Fibromyalgia | Endometriosis |
|---|---|---|---|
| Base population (EHR visits in both 2021–22 and 2022–23) | 262,790 | 262,790 | 262,790 |
| Recorded as female | – | – | 164,840 |
| Completed a health history survey (day 0) | 162,560 | 162,560 | 104,250 |
| Aged 18 years or older at day 0 | 162,560 | 162,560 | 104,250 |
| A visit in the 365 days before day 0 | 155,940 | 155,940 | 99,960 |
| No qualifying code by day 0 (analysis cohort) | 149,860 | 150,330 | 97,590 |
| of whom day 0 was before July 1, 2021 | 68,880 | 69,000 | 46,010 |

**Table S6: Sensitivity analyses: AUROC of the summary-count scores.** AUROC (95% CI) in the validation split. Main split: the split of Table 1 in the main text. All other rows use a second 80/20 split defined by a hash of the participant identifier, with every model refit. No later code: self-report among validation participants with no later code. Answer by day 30: self-report label limited to a first-told answer dated by day 30, self-report score refit. 730-day follow-up: validation participants with at least 730 days from day 0 to their last recorded visit. Common features, rounded counts: both scores fitted from counts rounded up to 10 of the features eligible for both labels. Baselines: a logistic regression on age, sex, and log visit counts trained on each label, and the number of symptom groups coded, which uses no label. Supplementary Methods defines each analysis. CI, confidence interval.

| Analysis | Target | Trained on self-report | Trained on later code |
|---|---|---|---|
| *Chronic fatigue* | | | |
| Main split (Table 1) | Uncoded self-reporters | 0.74 (0.72–0.75) | 0.67 (0.65–0.69) |
| Second split | Self-report | 0.75 (0.73–0.76) | 0.68 (0.67–0.70) |
| | Uncoded self-reporters | 0.74 (0.73–0.75) | 0.68 (0.66–0.69) |
| | Later code | 0.69 (0.67–0.71) | 0.70 (0.68–0.72) |
| No later code | Self-report | 0.74 (0.73–0.76) | 0.68 (0.66–0.69) |
| Answer by day 30 | Uncoded self-reporters | 0.74 (0.73–0.75) | 0.68 (0.66–0.69) |
| | Later code | 0.69 (0.67–0.71) | 0.70 (0.68–0.72) |
| 730-day follow-up | Uncoded within 730 days | 0.76 (0.74–0.78) | 0.71 (0.69–0.74) |
| | Later code within 730 days | 0.70 (0.68–0.74) | 0.71 (0.68–0.74) |
| Common features, rounded counts | Uncoded self-reporters | 0.74 (0.73–0.75) | 0.68 (0.66–0.69) |
| | Later code | 0.69 (0.67–0.70) | 0.70 (0.68–0.72) |
| Baseline: age, sex, visits | Uncoded self-reporters | 0.67 (0.65–0.68) | 0.64 (0.62–0.65) |
| Baseline: number of symptom groups | Uncoded self-reporters | 0.59 (0.58–0.61), untrained | |
| *Fibromyalgia* | | | |
| Main split (Table 1) | Uncoded self-reporters | 0.73 (0.71–0.74) | 0.64 (0.62–0.66) |
| Second split | Self-report | 0.75 (0.74–0.77) | 0.67 (0.65–0.69) |
| | Uncoded self-reporters | 0.75 (0.73–0.76) | 0.66 (0.64–0.67) |
| | Later code | 0.78 (0.75–0.80) | 0.78 (0.75–0.81) |
| No later code | Self-report | 0.75 (0.73–0.76) | 0.66 (0.64–0.67) |

**Table S6:** (continued)

| Analysis | Target | Trained on self-report | Trained on later code |
|---|---|---|---|
| Answer by day 30 | Uncoded self-reporters | 0.75 (0.73–0.76) | 0.66 (0.64–0.67) |
| | Later code | 0.78 (0.75–0.80) | 0.78 (0.75–0.81) |
| 730-day follow-up | Uncoded within 730 days | 0.73 (0.70–0.75) | 0.67 (0.64–0.69) |
| | Later code within 730 days | 0.79 (0.75–0.83) | 0.79 (0.75–0.83) |
| Common features, rounded counts | Uncoded self-reporters | 0.74 (0.73–0.76) | 0.66 (0.64–0.67) |
| | Later code | 0.77 (0.74–0.80) | 0.78 (0.75–0.81) |
| Baseline: age, sex, visits | Uncoded self-reporters | 0.67 (0.65–0.68) | 0.65 (0.64–0.67) |
| Baseline: number of symptom groups | Uncoded self-reporters | 0.48 (0.46–0.50), untrained | |
| *Endometriosis (female participants)* | | | |
| Main split (Table 1) | Uncoded self-reporters | 0.61 (0.60–0.62) | 0.53 (0.51–0.54) |
| Second split | Self-report | 0.60 (0.58–0.61) | 0.52 (0.51–0.53) |
| | Uncoded self-reporters | 0.60 (0.58–0.61) | 0.52 (0.50–0.53) |
| | Later code | 0.54 (0.50–0.59) | 0.72 (0.69–0.76) |
| No later code | Self-report | 0.60 (0.58–0.61) | 0.52 (0.50–0.53) |
| Answer by day 30 | Uncoded self-reporters | 0.60 (0.58–0.61) | 0.52 (0.50–0.53) |
| | Later code | 0.54 (0.50–0.59) | 0.72 (0.69–0.76) |
| 730-day follow-up | Uncoded within 730 days | 0.60 (0.58–0.62) | 0.52 (0.50–0.54) |
| | Later code within 730 days | 0.56 (0.49–0.63) | 0.73 (0.67–0.78) |
| Common features, rounded counts | Uncoded self-reporters | 0.60 (0.58–0.61) | 0.52 (0.50–0.53) |
| | Later code | 0.54 (0.50–0.58) | 0.73 (0.69–0.76) |
| Baseline: age, sex, visits | Uncoded self-reporters | 0.58 (0.56–0.59) | 0.44 (0.43–0.46) |
| Baseline: number of symptom groups | Uncoded self-reporters | 0.52 (0.51–0.54), untrained | |
| Women aged 18–44 | Uncoded self-reporters | 0.63 (0.60–0.67) | 0.53 (0.49–0.57) |
| | Later code | 0.63 (0.58–0.68) | 0.67 (0.63–0.72) |

**Table S7: AUROC for uncoded self-reporters by subgroup.** Second split, with every model refit. AUROC (95% CI) of the summary-count scores trained on self-report and on later codes. Counts are rounded up to the next multiple of 10. –: withheld so that a suppressed count cannot be recovered from the table. Dates are shifted by 1 to 365 days per participant, so the day 0 period is approximate. CI, confidence interval.

| Group | Participants | Uncoded self-reporters | Trained on self-report | Trained on later code |
|---|---|---|---|---|
| *Chronic fatigue* | | | | |
| All validation participants | 30,180 | 1,460 | 0.74 (0.73–0.75) | 0.68 (0.66–0.69) |
| Female | 19,220 | 1,180 | 0.73 (0.71–0.74) | 0.66 (0.65–0.68) |
| Male | 10,770 | – | 0.73 (0.70–0.76) | 0.69 (0.65–0.72) |
| Aged 18–44 | 7,440 | 520 | 0.76 (0.73–0.78) | 0.71 (0.69–0.73) |
| Aged 45–64 | 11,190 | 640 | 0.73 (0.71–0.75) | 0.67 (0.65–0.69) |
| Aged 65+ | 11,560 | 300 | 0.70 (0.67–0.73) | 0.66 (0.62–0.69) |
| Day 0 before July 1, 2021 | 13,990 | 610 | 0.76 (0.74–0.78) | 0.72 (0.69–0.74) |
| Day 0 on or after July 1, 2021 | 16,200 | 850 | 0.72 (0.71–0.74) | 0.65 (0.63–0.67) |
| *Fibromyalgia* | | | | |
| All validation participants | 30,300 | 870 | 0.75 (0.73–0.76) | 0.66 (0.64–0.67) |
| Female | 19,110 | 760 | 0.71 (0.69–0.73) | 0.61 (0.60–0.63) |
| Male | 11,000 | – | 0.75 (0.71–0.81) | 0.71 (0.66–0.77) |
| Aged 18–44 | 7,470 | 210 | 0.79 (0.76–0.82) | 0.68 (0.65–0.72) |
| Aged 45–64 | 11,140 | 420 | 0.74 (0.72–0.76) | 0.65 (0.63–0.68) |
| Aged 65+ | 11,690 | 240 | 0.69 (0.65–0.73) | 0.61 (0.58–0.65) |
| Day 0 before July 1, 2021 | 14,010 | 330 | 0.72 (0.70–0.75) | 0.67 (0.64–0.70) |
| Day 0 on or after July 1, 2021 | 16,290 | 540 | 0.76 (0.74–0.78) | 0.64 (0.63–0.66) |
| *Endometriosis (female participants)* | | | | |
| All validation participants | 19,660 | 1,600 | 0.60 (0.58–0.61) | 0.52 (0.50–0.53) |
| Aged 18–44 | 5,640 | 270 | 0.63 (0.60–0.67) | 0.53 (0.49–0.57) |
| Aged 45–64 | 7,790 | 740 | 0.57 (0.55–0.60) | 0.55 (0.52–0.57) |
| Aged 65+ | 6,240 | 590 | 0.54 (0.52–0.57) | 0.53 (0.51–0.55) |
| Day 0 before July 1, 2021 | 9,390 | 720 | 0.60 (0.59–0.62) | 0.52 (0.51–0.54) |
| Day 0 on or after July 1, 2021 | 10,280 | 880 | 0.59 (0.57–0.61) | 0.51 (0.49–0.53) |
